# Monitoring occurrence of SARS-CoV-2 in school populations: a wastewater-based approach

**DOI:** 10.1101/2021.03.25.21254231

**Authors:** Victor Castro Gutierrez, Francis Hassard, Milan Vu, Rodrigo Leitao, Beata Burczynska, Dirk Wildeboer, Isobel Stanton, Shadi Rahimzadeh, Gianluca Baio, Hemda Garelick, Jan Hofman, Barbara Kasprzyk-Hordern, Rachel Kwiatkowska, Azeem Majeed, Sally Priest, Jasmine Grimsley, Lian Lundy, Andrew C Singer, Mariachiara Di Cesare

## Abstract

Clinical testing of children in schools is challenging, with economic implications limiting its frequent use as a monitoring tool of the risks assumed by children and staff during the COVID-19 pandemic. Here, a wastewater based epidemiology approach has been used to monitor 16 schools (10 primary, 5 secondary and 1 post-16 and further education for a total of 17 sites) in England. A total of 296 samples over 9 weeks have been analysed for N1 and E genes using qPCR methods. Of the samples returned, 47.3% were positive for one or both genes with a frequency of detection in line with the respective community. WBE offers a promising low cost, non-invasive approach for supplementing clinical testing and can offer longitudinal insights that are impractical with traditional clinical testing.

## Introduction

The role of children and schools in the transmission of SARS-CoV-2 remains a matter of debate ^1,2,3^. Data suggest that children and adolescents (<18 years) account for less than 5% of the overall COVID-19 cases ^4,5^ and that this age group is characterised by milder or asymptomatic forms of the disease ^6^, different symptoms ^6^ and clinical outcomes from adults ^1,6,7^. However, children are susceptible to and can transmit SARS-CoV-2 within the school setting and from the school out to the community ^8^. Hence, there is a need for better methods to make schools safer environments and more covid-secure for children and staff. Testing will play a key role in this^9^.

Monitoring SARS-CoV-2 transmission and cases of COVID-19 in schools is challenging. Initial diagnostic testing focused on symptomatic patients, hence, increasing the likelihood that milder cases were missed, and no asymptomatic cases detected ^1^. To overcome the limits of symptoms-based testing, mass testing is currently used to identify positive cases in schools ^1^. While this approach would provide information on the burden of infection in schools, it is characterised by important organisational (e.g. delivery, parental consent) and economic burdens, putting in question its long-term sustainability as a primary surveillance system. Moreover, mass testing provides a picture of the status of the school population at a specific point in time, lending no insight into the days in between testing.

Wastewater-based epidemiology (WBE) is a promising non-invasive tool that can support the COVID-19 response as part of an early-warning system, providing data at a local community level to proactively inform public health care strategies and mitigate escalating demands on health care providers ^10,11,12^. SARS-CoV-2 has been identified in adult and child faeces and urine at different stages of the infection ^13^. A recent meta-analysis has estimated the mean duration time of SARS-CoV-2 shedding in stools to be 17.2 days [95% Confidence Interval 14.4–20.1], longer in duration than in any other body fluid ^14^. Wastewater is now widely used as a SARS-CoV-2 surveillance tool at sewer catchment level via the collection of grab and composite samples at wastewater treatment plant inlets ^11,15^. Near Source Tracking (NST)^16^ is conducted at a small sub-catchment scale (i.e., a building), permitting detection of small clusters or even individual COVID-19 cases in locations such as care homes^17^, hospitals ^18^, universities and student dormitories ^19,20^, aircraft ^10, 21^ and cruise ships ^10^. Here, we report on the first ever study that has explored the use of a WBE approach to identify the presence of SARS-CoV-2 in primary and secondary school wastewater, in England (UK).

## Methods

Sixteen schools (10 primary, 5 secondary and 1 post-16 and further education for a total of 17 sites) in England have taken part in the School wasTEwater-based epidemiological suRveillance systeM for the rapid identification of COVID-19 outbreaks (TERM) study. Schools were located across neighborhoods with different levels of deprivation and with diverse school populations: two in South West England, one in North West and one in South East England (**Table 1**).

**Table 1.** Schools characteristics

| **Phase of education** | **Frequency of schools** |
| --- | --- |
| Primary | 10 |
| Secondary | 5 |
| Post 16 | 1 |
| **LSOAs quintiles (IMD)** |  |
| Q1 - lower | 7 |
| Q2 | 3 |
| Q3 | 3 |
| Q4 | 2 |
| Q5 – higher | 1 |
| **Pupils classified as white British** |  |
| Very Low: Up to 20% | 5 |
| Low: Between 21% and 40% | 1 |
| Med: Between 41% and 60% | 2 |
| High: Between 61% and 80% | 4 |
| Very High: Between 81% and 100% | 3 |
| NA | 1 |
|  | **Min-Max** |
| **Number of pupils (official)** | 143-2061 |
| **% pupils using free school meals** | 0-39.8 |
| **% of pupils classified as white British ethnic origin** | 1.2-95.1 |
| **IMD Rank out of LSOAs (32,844)** | 890-27291 |
| **IMD Rank out of LA (317)** | 36-238 |
| **Rate per 100,000 population (cum)** | 209.74-1020.37 |
| **Rate per 100,000 population (weekly)** | 15.09-122.06 |

Note: NA not available; rates per 100,000 population (both cumulative and weekly); IMD: Index of Multiple Deprivation; LSOAs: Lower Layer Super Output Area <sup>34,35</sup>

## Sampling Collection

Sampling began on 20th October 2020, approximately six weeks after the start of the 2020-2021 school year. Composite wastewater samples were initially collected twice a week (Tuesday and Thursday; from 8 am to 3 pm) as 7-hour time-proportional composites at a sampling frequency of 60 seconds ‘on’, followed by 4 minutes, ‘off’ using an Aquacell P2-COMPACT (Aquamatic) autosampler. The maximum pumping rate in cases where a steady flow of wastewater was present was approximately 50 ml/min. After an initial trial period, a second autosampler (Aquacell P2-COMPACT (Aquamatic)) was installed at 7 locations (in 6 schools), to collect samples at a frequency of one minute on/ one minute off between 12pm and 2pm. The purpose of this was to assess whether increasing the frequency of sampling over the lunchtime period (when we assume there is greater opportunity to use the bathroom) improved the likelihood of detecting SARS-CoV-2. In addition, from the 4th November 2020, the frequency of sampling was increased to 4 days per week (Monday to Thursday). At the end of each school day, one litre from the maximum of 5 litres of wastewater collected in a plastic container was decanted from each autosampler, after thorough mixing, into a separate plastic (polypropylene or polyethylene terephthalate) sample bottle and immediately couriered to the laboratory on melting ice. Sample temperature was checked on receipt in the laboratory. Storage temperatures were monitored daily to ensure stable temperature in the range of 2.5 – 4.0°C. Aliquots of these samples underwent RNA extraction and cryogenic sample preservation (−80°C) within 24 hours of sample receipt at the laboratory. An overview of sample collection data is provided in Table 2.

**Table 2.** Overview of school locations, sampling dates, collection volumes and ragging (other) incidents that prevented sample collection

| **Area (number of schools; number of autosamplers)** | **Sample collection dates (start date - end date)** | **Total number of days of collection** | **Volume collected** | **Number of ragging (or other issues<sup>1</sup>) incidents preventing sample collection** |
| --- | --- | --- | --- | --- |
| Area 1 (4; 6) | 20/10/20 - 17/12/20 | 83 | 0-5L | 2 (7) |
| Area 2 (6; 10) | 04/11/20 - 17/12/20 | 114 | 0-5L | 3 (8) |
| Area 3 (4; 5) | 17/11/20 - 17/12/20 | 69 | 0-5L | 0 (1) |
| Area 4 (3; 3) | 10/12/20 - 17/12/20 | 14 | 0.5-5L | 0 |

Key: <sup>1</sup> no sample collected for variety of reasons including ragging (number not in brackets) or due to either battery failure, in pipe blockages, autosampler tube out of alignment, unable to access site/inset day or reason not given (summed figure in brackets)

## Protocol for sample analysis

Each wastewater sample was analysed in the laboratory for total suspended solids, ammonium (NH_4_-N), orthophosphate (PO_4_-P), total chemical oxygen demand (tCOD), soluble chemical oxygen demand (sCOD), pH, conductivity and dissolved oxygen according to standard methods for the examination of water and wastewater ^22^. The method used for SARS-CoV-2 RNA analysis is described in detail in a substantive paper ^23^. In summary, school wastewater samples were centrifuged (30 minutes at 3,000 x g at 4°C) and supernatants were spiked with an extraction control murine norovirus before concentration using the poly-ethylene glycol (PEG) precipitation method with an overnight incubation ^24^. A final concentrate was obtained by centrifugation (10,000 × g for 30 min at 4°C), the PEG was removed by pouring, followed by a further centrifugation step / PEG removal step (10,000 × g for 10 min at 4°C) and the resulting pellet resuspended in 0.5 mL of molecular biology grade phosphate buffered saline (PBS), pH 7.4. Viral extraction from wastewater concentrates was carried out using the NUCLISENS^®^ RNA extraction kit on a MINIMAG^®^ (BioMérieux, France). SARS-CoV-2 RNA detection was performed by RT-qPCR using the RNA UltraSense™ One-Step Quantitative RT-PCR System (ThermoFisher, UK) targeting the nucleoprotein (N), N1 fragment ^25^, and envelope protein (E) gene ^26^ using a QuantStudio™ 7 Pro Real-Time PCR System (ThermoFisher, UK). RNA samples were analysed in duplicate alongside negative (nuclease-free water) controls. The RNA extracts were quantified by plotting the quantification cycles (CT) to an external standard curve constructed from commercially available synthesised plasmids containing the target sequence. The empirical limit of quantification (LOD) was calculated based on a method outlined previously^23^ and denoted 1268 GC / L for N1 and 2968 GC / L for E. The Limit of Quantification (LOQ) was 9196 GC / L for N1 and 21300 GC / L for E. This was determined through spiking SARS-CoV-2 negative RNA extracts from school wastewater with a range of defined quantities of Armored RNA standard (Asuragen Quant SARS-CoV-2 Panel - 52036, VH Bio Ltd., UK) with the LOQ being the lowest concentration which achieved a CV value not exceeding 25%. During the study, a positive detection was considered when a sample exceeded LOD alongside no significant amplification in the negative control. In a pilot study based on duplicate extractions from 37 different wastewaters using this method. For samples which we could positively quantify, the coefficient of variation (CV) reported as the average CV (minimum – maximum) for full method replicates including concentration, RNA extraction and RT-qPCR for N1 was 9.3% (0.42-23.8)% in wastewaters ranging from LOQ-8 × 10^6^ GC / L. The CV for E was 14.5% (0.3-34 %) in wastewaters ranging from LOQ-9.38 × 10^6^ GC / L.

## Results

A total of 296 samples were analysed for both standard wastewater parameters and SARS-CoV-2 RNA. In terms of wastewater characteristics, concentrations determined were highly heterogeneous (see **Appendix Table 1** for data for all samples and for samples in which the N1 and/or E gene were detected). Whist median values for all parameters are slightly lower in samples in which a positive signal was detected, the reported range for several parameters indicate that SARS-CoV-2 RNA can be detected in school-derived wastewater samples even when, for example, TSS and NH_4_-N concentrations are four orders of magnitude below reported medians, an indication that few individuals have contributed waste products to the sample. 47.3% of the 296 samples collected returned positive for one or both genes. 129 (43.6%) samples were positive for the N1 gene and 75 (25.3%) samples were positive for the E gene. Sixty-three samples were positive for both N1 and E genes (21.3%), 65 samples were positive for the N1 gene but not the E gene (22.0%) and 12 samples (4.0%) were positive for the E gene but not the N1 gene (**Figure 1 and Figure 2**). In 42.1% of samples, a first positive detection for both genes was predated by a positive detection of one targeted gene and in 36.8% by a non-detection (in 21.1% of cases no sample was collected on the sampling day before the positive case). 80.4% of the samples collected during the first week of December returned positive. This percentage remained around 56% and 51% during the second and third week of December (**Table 3**). At the beginning of the study period the rate of positivity in samples collected in primary schools was higher than those observed in secondary schools. From week 5 (last week of November) the rate of positivity was consistently higher in samples from secondary schools settings when compared to primary schools, with a positive rate of 88.9% in the first week of December. Levels of GC/L varies between 1333 GC/L and 1.68*10^6^ GC/L for the N1 gene target, and between 3067 GC/L and 1.31*10^6^ GC/L for the E gene target. The minimum Ct value was 31.0 for the N1 gene and 28.2 for the E gene (**Figure 3**).

**Table 3.**
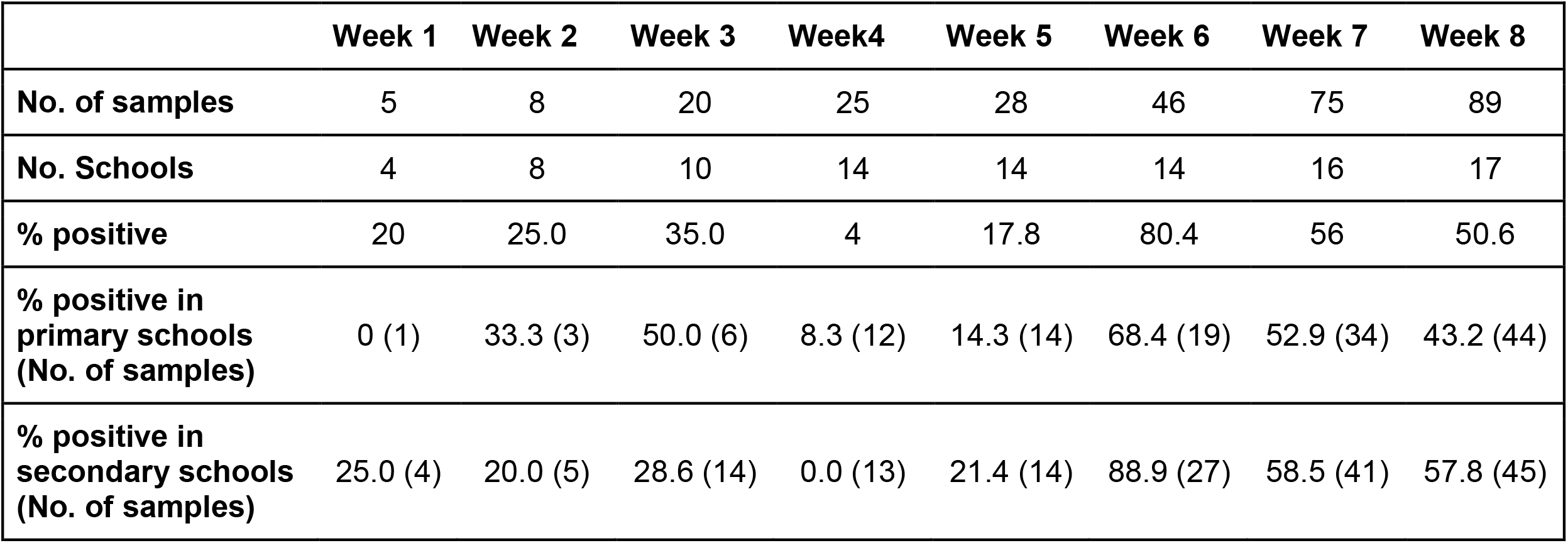
Number of samples, percentage of positive cases (total, and by stage of education) by week

**Figure 1.**
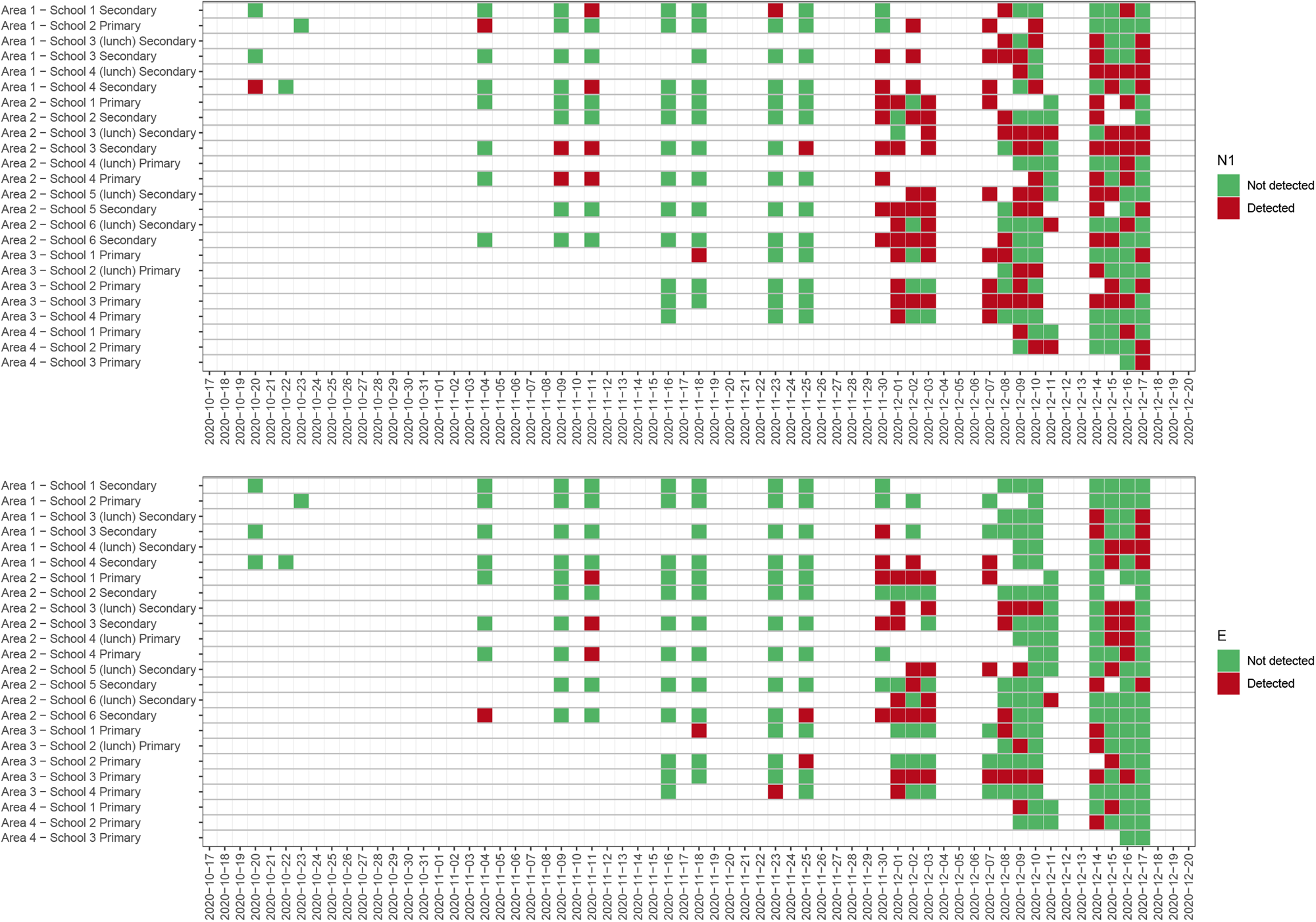
Heatmap detection/non-detection for N1 gene (a) and E gene (b)

**Figure 2.**
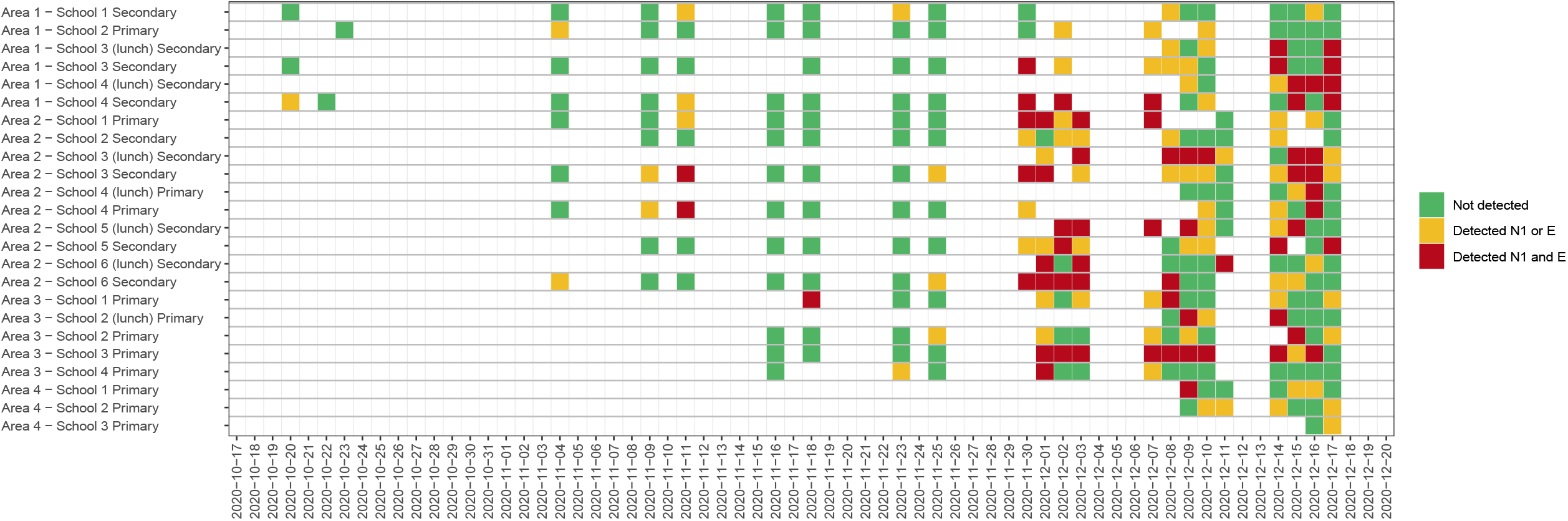
Heatmap detection/non-detection N1 gene and E gene combination

**Figure 3.**
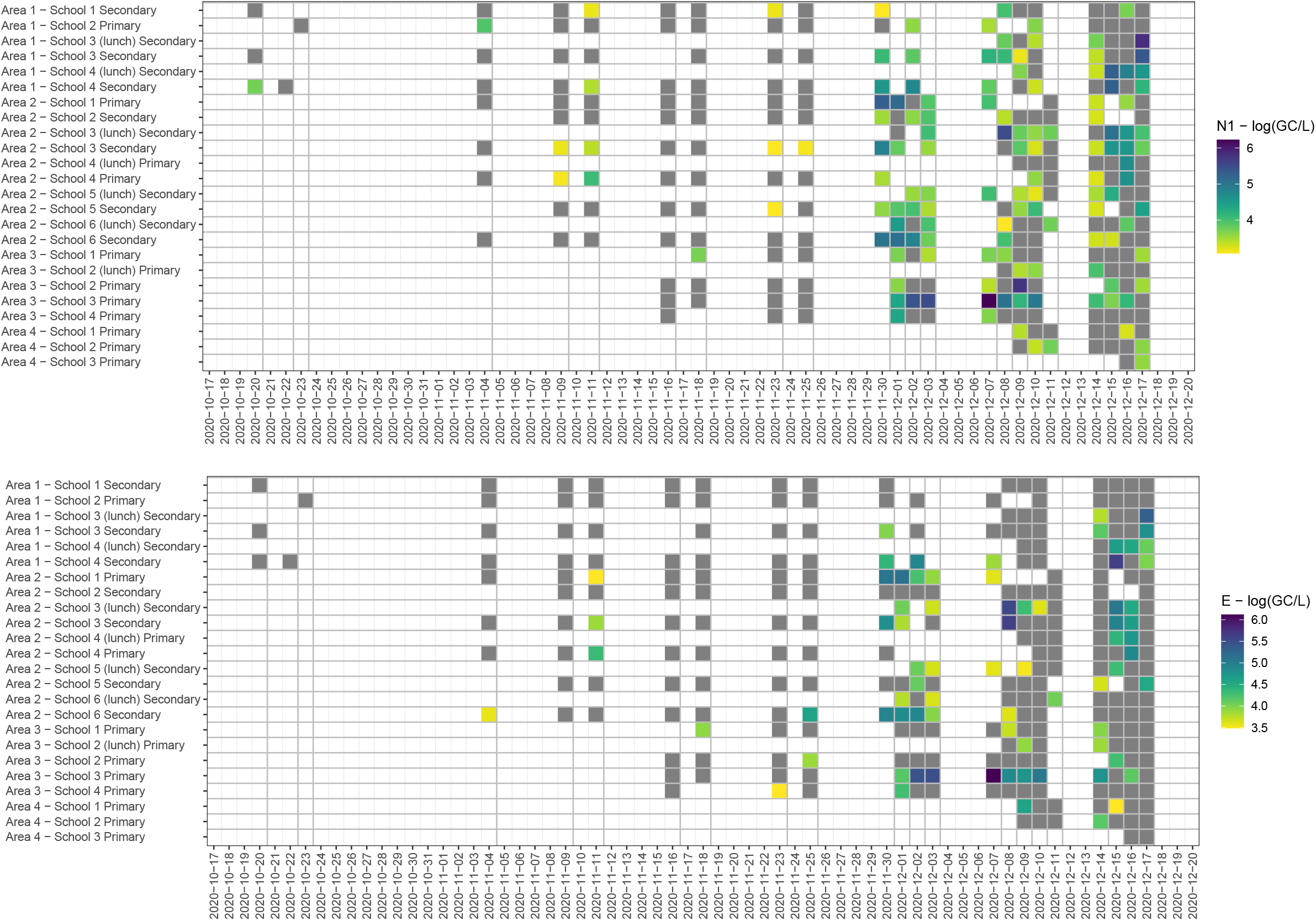
Heatmap daily log10(GC/L) N1 gene and E gene (grey indicated non detection)

In Figure 4, weekly new COVID-19 cases in the community (reported for each school’s Middle Layer Super Output Area (MSOA) and all the adjacent MSOAs, for a total of 92 MSOAs) are presented together with the percentage of positive samples collected each week. The increase/decrease in the frequency of detection of targeted genes are in line with the increase/decrease in new cases with a lag of around 14 days.

**Figure 4.**
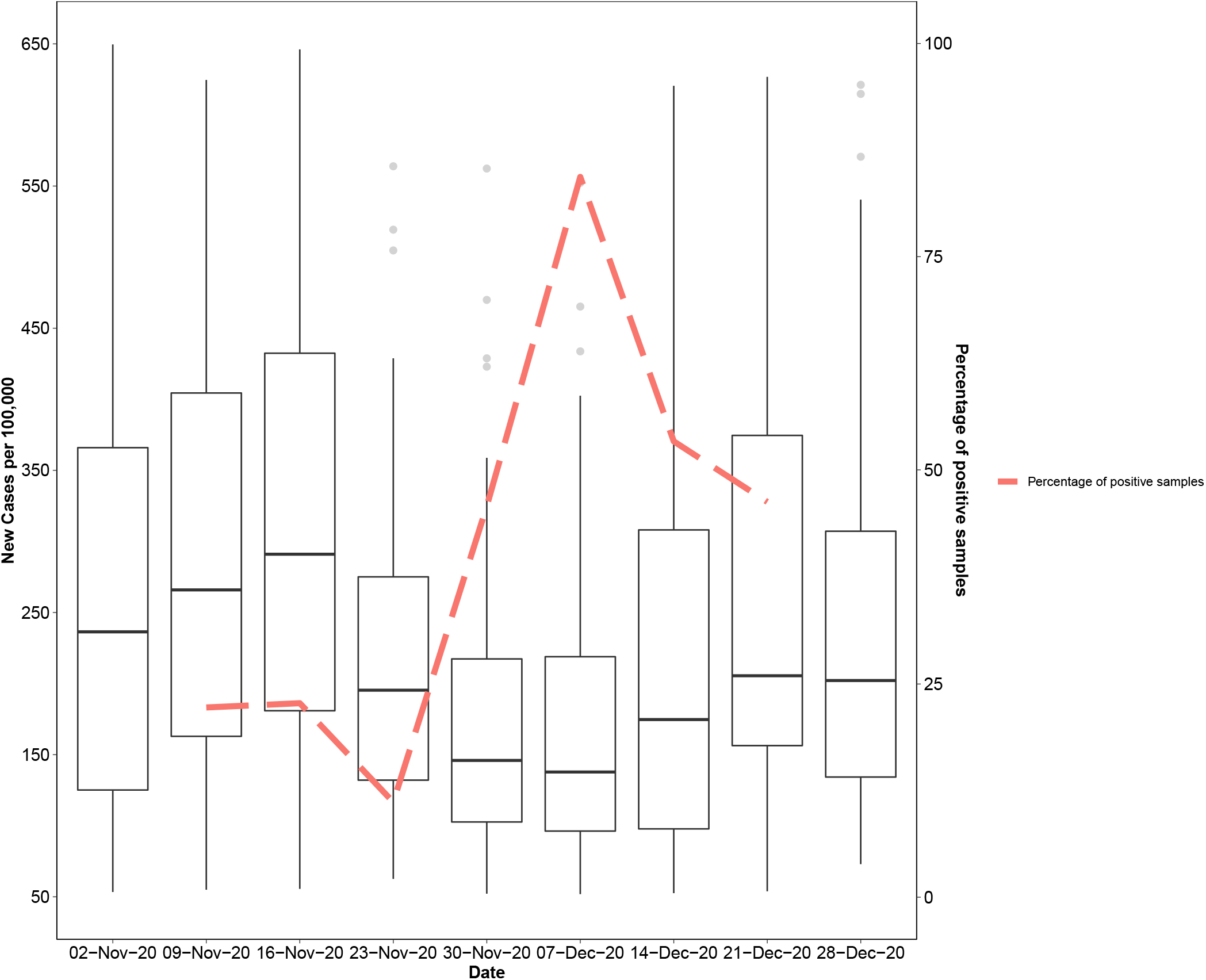
Middle Layer Super Output Areas (MSOAs) COVID-19 new cases per 100,000 by week and percentage of positive school samples. Note: Each box shows the distribution of new cases per 100,000 across the MSOA* of each school and all the adjacent MSOAs (92 MSOAs). Solid lines show medians, the boxes show Inter-Quartile Ranges, the whiskers show ranges. Dashed line show the percentage of weekly positive samples (weeks in this figure are defined based on the MSOA data availability and may differ from the weeks of the study). * MSOAs are a geographic hierarchy designed to improve the reporting of small area statistics in England and Wales Source: MSOA level data extracted from https://coronavirus.data.gov.uk/details/download

Among both all-day and lunch samples collected in the same school (52 times), 61.5% of these both samples returned consistently positive (or negative); in 23.1% of the occasions, all-day samples were positive (but not the lunchtime) while in 15.4%, the lunchtime samples were positive (but not the all-day).

## Discussion

This is the first published study to report on the use of Near Source Tracking (NST) for the detection of SARS-CoV-2 in primary and secondary schools. Data suggest that the virus was circulating in schools during the autumn and early winter, and that the frequency of detection intensified in line with the increase in new cases notifications in the community. Data collected confirm that despite the episodic nature of wastewater flows in schools, presence of SARS-CoV-2 within the school setting can be identified, which suggests that the use of wastewater epidemiology NST in schools can play a valuable role in monitoring and motivating rapid action. The implications of these findings reinforce the need for continuous monitoring of the wastewater, especially when employing NST with its inherent episodic flow.

Viral material detected in samples has two possible origins: shedding from students and/or staff and/or visitors. Validating the source of viral shedding requires regular asymptomatic testing of students as well as staff; during the period of analysis this was not the strategy in English schools, though there are plans to test secondary school students going forward ^28^.

Few, if any, data are available on the toilet use habits of either primary or secondary school-aged children. However, the anecdotal assumption/hypothesis that students and staff only reluctantly use toilets at schools, is incorrect. The data clearly indicate that sufficient numbers of attendees of school are contributing to the waste stream to yield upwards of 52% positive samples for SARS-CoV-2 over the period of analysis. Those with mild symptoms or are asymptomatic for COVID-19 may be more likely to defecate than uninfected students. Some evidence to support loose bowel movements is emerging in the literature ^29^, with 19% of children who tested positive reporting gastrointestinal symptoms including diarrhoea ^30^. However, it remains unknown the extent to which the frequency of bowel movements increases in mild or asymptomatic children with COVID-19. SARS-CoV-2 in urine, vomit, handwashing and/or mucus (from nose blowing) cannot be ruled out as a source of some positive detects in the wastewater.

Surveillance data from NST were used to confirm the presence or absence of SARS-CoV-2 in the wastewater. Although the data are quantitative, it is not currently possible to identify the number of unique flush events represented within a composite sample. It is tempting to normalise the data to the population, for example to estimate the proportion of the sample population that are infected, but the number of individuals who have had a bowel movement on a given day remains unknown. Yet, the intervention should not be dependent on the number of infected people, with mass testing performed to identify infected individuals and support their isolation. Given the uncertainty around translating wastewater concentrations of SARS-CoV-2 into counts of infected individuals, NST is, at this stage, better employed as an early warning tool, by signaling presence of the virus, than as a measure of transmission. Further work (i.e. introduction of continuous sampling, better understanding of viral shredding and robust modelling approaches) is required to enable estimation of the number of infected individuals.

There is still some uncertainty regarding the utility of qualitative measurements (≥ LOD) vs quantitative measurements (≥ LOQ) considering uncertainties resulting from the sampling and analysis. As a surveillance tool, the primary function is to find a pathogen in locations that it should not be, i.e., a school. Hence, use of the LOD (presence/absence) is perhaps justified as it demonstrates the main analyte is present in the sampled population. However, a school sample that reports <LOD is still at risk of false negatives given the limitations of the autosampler and its sampling schedule linked with the probability of capturing stool from all infected individuals (i.e. even a composite sample is only a fraction of the flow over a given time period). To illustrate this issue, it is currently likely that the autosampler only turns on just after the flush of an infected individual has passed by and been missed. In this scenario, one might expect either no or very low levels of SARS-CoV-2 in the autosampler reservoir, yielding a detectable (i.e. >LOD) but not quantifiable (<LOQ) result. In particular, if both the probability of toilet use in school and the probability of an individual in the school to be infected are considered together with the probability of the autosampler to capture stool from an infected individual (considering that only 20% of the flow is sub-sampled) it is evident that the likelihood of a positive detection is extremely low. To illustrate: an optimistic scenario of 50% of flushes containing faecal material, 1% positive cases in the school population, and 20% of wastewater sampled yields a probability of detecting an infected individual’s stool at <0.1%. Yet, here we report detection in approximately 50% of the samples. The apparent deviation from our assumptions suggests that some of the assumptions behind toilet behaviours in schools, faecal shedding among asymptomatic/pre-symptomatic individuals, and prevalence of gastrointestinal symptoms among infected individuals need to be researched further.

The addition of a second autosampler with a higher sampling rate positioned during the lunch periods revealed that defecation by infected individuals was spread throughout the day; with 23.1% of SARS-CoV-2 positive samples measured from the full day autosampler only (i.e. not detected in the lunch-specific sampler). Illustrative of the importance of high sampling rates when conducting NST, the lunchtime sampler yielded SARS-CoV-2 positive samples several times without any detection in the full day sampler (15.4%). Each flush event is an important source of data and each missed bowel movement risks a false negative result, i.e a school could be characterised as free from SARS-CoV-2 when in fact it contains infected staff/ students. Above discussion warrants further study into the design of new continuous autosamplers, sensing approaches and passive sampling solutions. Novel sensors might one day be deployable which would allow for continuous measurements, in real-time. Further work is needed to integrate whole-genome sequence analysis into NST, which could provide invaluable information on the extent of SARS-CoV-2 transmission in the community and insights into new variants of concern.

Currently best practices and specific guidance on how local health protection teams can respond to NST WBE data is lacking. An approach is needed to co-produce with stakeholders protocols to integrate these data into existing practices which will define when, how and to whom the data should be shared. The public health benefits must, however, be constantly weighed against ethical trade-offs since human waste products contain a wealth of information about behaviours as well as health status, and at the level of a single building, it is difficult to preserve donor anonymity. As well as potentially infringing on individual autonomy, this raises the risk of stigmatising businesses, communities, or individuals with associated financial and social disbenefits. NST methods and tools are in development to support ethical sampling strategies, and robust and transparent reporting to inform public health action ^31,32^.

Despite its limitations, the data support the use of wastewater epidemiology NST in schools as a tool to trigger outbreak investigation or asymptomatic testing. This is an approach that could be used to identify new outbreaks, showing long term sustainability of WBE to enhance resilience in public health response in schools and other vulnerable or under sampled populations. Communication of positive signals within a school community represents the opportunity to develop a targeted approach. Definitive knowledge of the presence of SARS-CoV-2 in a school community permits stronger communication messaging about the need for mass testing of individuals and can help overcome barriers of risk denial/displacement and the commonly observed testing hesitancy. Results from the first round of the Schools Infection Survey (ONS) suggest that the participation rate among pupils was on average 17% (51% among staff) ^33^; highlighting a critical need to encourage the increased uptake of mass testing amongst school pupils and their families. In addition, regular feedback of results to each school and its wider community may improve engagement with non-pharmaceutical interventions and implementation of infection prevention and control measures such as decreasing bubble sizes, staggering start/ finish/ break times and enhanced cleaning protocols. Finally, WBE could provide valuable insights on the local epidemiology of COVID-19 into areas/schools with very low uptake of school mass testing and high level of hesitancy to vaccination.

## Data Availability

Anonymised data that support the findings of this study are available on request from the corresponding author (MDC).

## Contributors

MDC, LL, FH, AS designed the study. FH led the lab analysis with the support of AS, VCG, RL, MV, BB, DW, IS. MDC, LL, SR prepared the results. MDC, LL, FH, AS wrote the first draft of the paper. MDC oversaw the work. All other authors contributed to the final version. All authors read and approved the final version of the manuscript.

## Disclaimer

JG - The views expressed in this paper are those of the author and do not necessarily reflect the views or policies of the Department of Health and Social Care.

## Code availability

Not applicable

## Acknowledgment

We thank Prof. Davey Jones and Dr Kata Farkas, Bangor University for providing MNV.

## Ethics

Ethical approval for this study was obtained from Middlesex University Ethic Committee (14795).

## Role of the funding source

The funder supported the identification of priority areas. The funder played no role in study design, data collection, data analysis, data interpretation, or writing of the report. The corresponding author had final responsibility for the decision to submit for publication.

## Funding

Department of Health and Social Care, UK

## Appendix

**Table 1.**
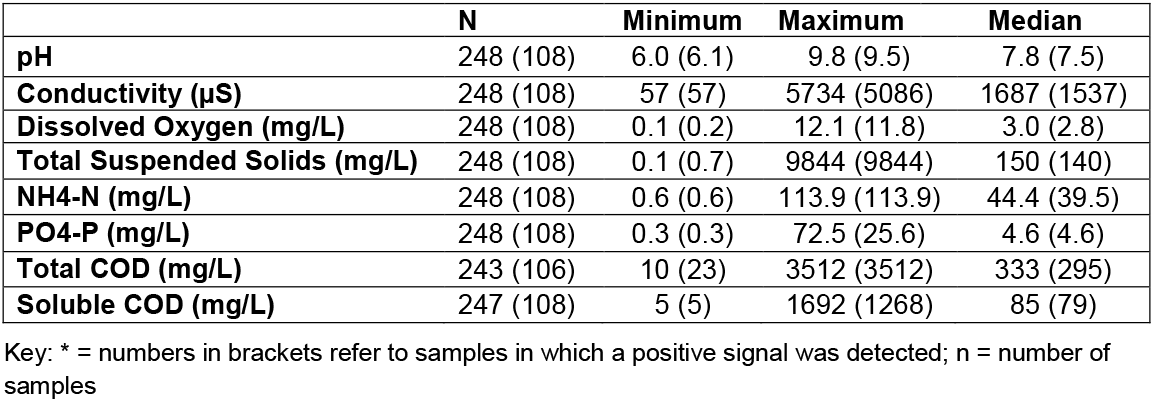
Overview of key wastewater characteristics in samples collected from all schools and in samples where the N1 and/or E gene were detected*

|  | **N** | **Minimum** | **Maximum** | **Median** |
| --- | --- | --- | --- | --- |
| **pH** | 248 (108) | 6.0 (6.1) | 9.8 (9.5) | 7.8 (7.5) |
| **Conductivity (µS)** | 248 (108) | 57 (57) | 5734 (5086) | 1687 (1537) |
| **Dissolved Oxygen (mg/L)** | 248 (108) | 0.1 (0.2) | 12.1 (11.8) | 3.0 (2.8) |
| **Total Suspended Solids (mg/L)** | 248 (108) | 0.1 (0.7) | 9844 (9844) | 150 (140) |
| **NH<sub>4</sub>-N (mg/L)** | 248 (108) | 0.6 (0.6) | 113.9 (113.9) | 44.4 (39.5) |
| **PO<sub>4</sub>-P (mg/L)** | 248 (108) | 0.3 (0.3) | 72.5 (25.6) | 4.6 (4.6) |
| **Total COD (mg/L)** | 243 (106) | 10 (23) | 3512 (3512) | 333 (295) |
| **Soluble COD (mg/L)** | 247 (108) | 5 (5) | 1692 (1268) | 85 (79) |

Key: \* = numbers in brackets refer to samples in which a positive signal was detected; n = number of samples

## Notes

### Competing Interest Statement

The authors have declared no competing interest.

### Funding Statement

Funding:
Department of Health and Social Care, UK
Role of the funding source:
The founder supported the identification of priority areas. The founder played no role in study design, data collection, data analysis, data interpretation, or writing of the report. The corresponding author had final responsibility for the decision to submit for publication.

### Author Declarations

Ethical approval for this study was obtained from Middlesex University Ethic Committee (14795).

